# Loss of CHGA protein as a potential biomarker for colon cancer diagnosis: a study on biomarker discovery by machine learning and confirmation by immunohistochemistry in colorectal cancer tissue microarrays

**DOI:** 10.1101/2022.04.04.22271362

**Authors:** Xueli Zhang, Hong Zhang, Chuanwen Fan, Bairong Shen, Xiao-Feng Sun

## Abstract

**Background:** Incidences of colon and rectal cancers have been parallelly and constantly increased, however, the mortality has been slightly decreased due to early diagnosis and better therapies during the last decades. Precise early diagnosis for colorectal cancer has been a great challenge in order to win chances for the best choices of cancer therapies.

**Patients and methods:** We have started with searching protein biomarkers based on our colorectal biomarker database (CBD), finding differential expressed genes (GEGs) and non-DEGs from RNA sequencing (RNA-seq) data, and further predicted new biomarkers on protein-protein interaction (PPI) networks by machine learning (ML) methods. The best-selected biomarker was further verified by receiver operating characteristic (ROC) test from microarray and RNA-seq data, biological network and functional analysis, and immunohistochemistry in the tissue arrays from 198 specimens.

**Results:** There were twelve proteins (MYO5A, CHGA, MAPK13, VDAC1, CCNA2, YWHAZ, CDK5, GNB3, CAMK2G, MAPK10, SDC2, and ADCY5) which were predicted by ML as colon cancer candidate diagnosis biomarkers. These predicted biomarkers showed close relationships with reported biomarkers on PPI network and shared some pathways. ROC test showed CHGA protein with the best diagnostic accuracy (AUC=0.9 in microarray data and 0.995 in RNA-seq data) among these candidate protein biomarkers. Furthermore, CHGA performed well in the immunohistochemistry test.

**Conclusions:** Protein expression of CHGA in the normal colorectal mucosa was lost in the colorectal cancers and the lose of CHGA protein might be a potential candidate biomarker for colon or even colorectal cancer diagnosis.

**Topic:** CHGA expression, colon cancer diagnosis

**Key Message:** The results of this study suggest that lose of CHGA expression from the normal colon and adjacent mucosa to colon cancer may be used as a valuable biomarker for early diagnosis of colon adenocarcinoma

## Introduction

Colon cancer contributes essentially to cancer mortality and morbidity [1]. In 2020, there will be 104610 new colon cancer cases and 53200 deaths caused by colon cancer in the United States, estimated by the National cancer institute [2]. Surgery is the primary treatment for early-stage colon cancer [3]. With the development of modern medicine and surgery technology, the 5-year survival rate of stage I and II has increased to more than 90% [4]. However, the rate of stage IV is around 10% [4]. What’s more, more than 50% of patients are already at late-stage colon cancer when they are diagnosed [5]. As such, the timely and accurate early diagnosis of colon cancer is highly needed.

Biomarkers are biological indicators for special clinical conditions or states, which have been reported many times, improving the diagnosis of colon cancer [6, 7]. In the previous work, our research group has established an integrated colorectal biomarker database (CBD), which has collected all the colon cancer-related biomarkers [7]. However, few of these biomarker has been used in clinical, and the effects are not convincing [7, 8]. Hence, It is needed to predict new biomarkers. Recently more and more studies have convinced that combining different single biomarkers together as multiple biomarkers could reach better clinical performance than single biomarkers [9-11]. Therefore, the development of multiple biomarkers could be a new direction in biomarker discovery.

Colon cancer and rectal cancer have many similar features in both genotype and phenotype, which are always grouped as colorectal cancer (CRC) [12]. It has been convinced that colon cancer and rectal cancer share many biomarkers [7]. Hence, the application of new colon cancer biomarkers in rectal cancer can be expected. The development of cancer is a continuous process. Many studies report that some diagnosis biomarkers can also serve as prognosis biomarkers in CRC [13, 14], which are considered as multiple-functional biomarkers. As such, the expansion of novel diagnosis biomarkers in prognosis is reasonable.

Network topology analysis is an important component of system biology study [15]. Many researches have proved that biomarkers occupy specific positions on the biological interaction networks [16-18]. Based on this theory, we predicted three novel miRNA biomarkers for colorectal cancer diagnosis, using network topology features from the miRNA-mRNA interaction network, and they showed good diagnosis value in the verification test by meta-analysis [18]. Proteins are the major part of colon cancer biomarkers [7]. The String database contains highly credible human protein-protein interaction (PPI) networks collected from different resources, which could be the effective data source for protein-related network topology analysis [19].

Machine learning (ML) has been applied in bioinformatics and complex network analysis for many years [20]. Support Vector Machine (SVM) is a supervised based ML method focusing on classification and regression analysis, which has been developed as a popular method in bioinformatics since its good accuracy and robustness [21]. Several published studies utilized SVM and PPI networks in cancer biomarker prediction [22-24]. However, none of them used identified biomarkers for the training dataset [22-24], which we think decreased the prediction credibility With its high robustness, low heterogeneity, and extensive adaptability, Bioinformatics (dry lab) experiments have become a new focus in the biomedicine field, especially in cancer biomarker discovery [25]. Traditional biomedicine (wet lab) experiments are closer to the actual situation, which are convinced with high credibility. In the past years, our research groups predicted and identified several useful CRC biomarkers using traditional biomedicine experiments or bioinformatics [17, 18, 26, 27].

Chromogranin A or parathyroid secretory protein 1 (gene name CHGA) is a member of the grain family of neuroendocrine secretory proteins, and it is located in secretory vesicles of neurons and endocrine cells such as islet beta-cell secretory granules in the pancreas [28]. In humans, chromogranin A protein is encoded by the CHGA gene. In this study, we used the reported colon cancer diagnostic biomarkers to predict new biomarkers via ML methods, based on the topology features from PPI network. Diagnostic receiver operating characteristic (ROC) test, immunohistochemistry (IHC), and biological network and function analysis were conducted to make the verification and confirmed that CHGA could be a future biomarker for colon cancer diagnosis. Meanwhile, the multiple biomarkers consisting by the 12 predicted biomarkers have been convinced with high diagnostic accuracy. Further, the diagnosis and prognosis value of CHGA in both colon and rectal cancer were evaluated, which indicated that CHGA could be a promising diagnosis but not a prognosis biomarker in CRC.

## Methods and Methods

### Patients information

There are 198 specimens in the present study, including 55 primary tumours, 22 biopsies, 22 metastatic lymph nodes, 46 adjacent normal mucosae (adjacent to the primary tumor on the same histologic section) and 53 distant normal mucosae from the proximal or distal margin (4-35 cm from the primary tumor) of the resected colorectum. All the cases in the study were patients from the Southeast Swedish Health Care region. The detailed characteristics of samples are presented in Table 1. All the samples including adjacent normal mucosa, distant normal mucosa, primary cancers, and metastatic lymph nodes were paraffin-embedded and fabricated into TMA as the previous description [29]. The study was conducted in accordance with the Declaration of Helsinki, and the protocol was approved by the institutional review board of Linköping University, Sweden (Dnr-2012-107-31, Dnr 2014-79-31).

**Table 1.**
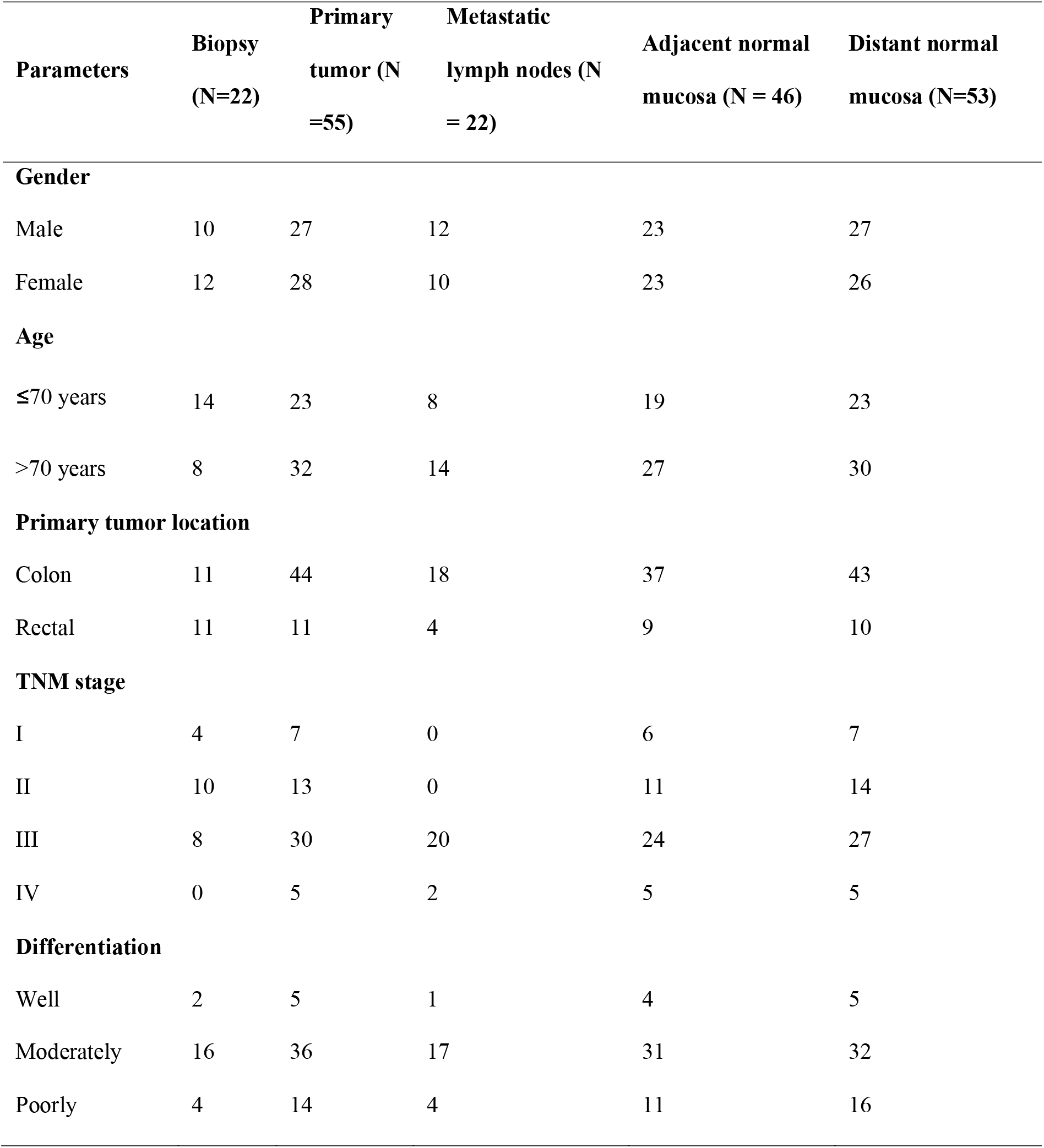
Characteristics of patients included in the present study

### Immunohistochemical assay and measurements of ChgA expression by IHC

IHC staining for ChgA expression was done as described in previous work on 5-μm TMA sections from paraffin-embedded TMA slides [29]. Briefly, the sections were deparaffinized, rehydrated and masked epitope retrieval. Then, after blocking the activity of endogenous peroxidase, the sections were incubated with the ChgA monoclonal rabbit anti-Human IgG (CM10C, BIOCARE MEDICAL) in a 1:100 dilution with antibody dilution buffer overnight. The next day, the sections were washed in PBS and then incubated with Envision System Labelled Polymer-HRP Anti-Rabbit (Dakocytomation) for 30 min. Next, the sections were subjected to 3,3’-diaminobenzidine tetrahydrochloride for 8 min and then counterstained with hematoxylin. Negative and positive controls were added in each staining run. All slides were measured by two independent investigators. ChgA stains were scored as previous description: 0, no staining; +1, < 2% staining in the normal intestinal or tumor cell; and +2, >2% staining in the normal intestinal or tumor cell [30].

### Data collection

We downloaded the Colon cancer differential expression (DE) data from the GEPIA (Gene Expression Profiling Interactive Analysis) database (http://gepia.cancer-pku.cn/index.html), which concluded normalized and comprehensive high-throughput RNA sequencing (RNA-Seq) data from the Cancer Genome Atlas (TCGA) and Genotype-Tissue Expression (GTEx) database. Two hundred seventy-five colon cancer patients and 349 normal controls were included. Linear model and the empirical Bayes method were used to calculate the DE genes (DEGs) by the limma package in R. P-value<0.05, and |Log_2_FC|>1 was selected as the cut off for the DEGs. All the DEGs, along with their statistics results, can be found in **Supplementary Material Table S1**.

The Human PPI network (confidence>0.7) was downloaded from the String database via the NDEx public server. Colon cancer diagnostic protein biomarkers were downloaded from the CBD database. **(Supplementary Material Table S2)**

Gene Expression Omnibus (GEO) database provided the microarray data named “GSE 44861” for verification of candidate biomarkers, which contained 111 colon tissues from tumors and adjacent noncancerous tissues. These GE data were from GPL3921 Platform.

### Colon cancer specific PPI network construction

The colon cancer DEGs were transferred to protein by searching in the NCBI protein database then mapped to the Human PPI network. The greedy search algorithm of jActiveModules in Cytoscape was used to find the most highly scored subnetwork from the human PPI network according to colon cancer patients DE genes’ p-value. In the greedy searching, firstly every p-value of DE gene will be transferred to z-score using the Stouffer’s Z-score model: less p value will have bigger z-score. Then a k-subnetwork will be given a z(A):

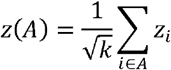

We selected the subnetwork with the highest summary z (A) after several iterations. This subnetwork was constructed by every highly scored DE genes along with one of its neighbouring genes. Here we named this subnetwork as Colon cancer specific PPI network (CCS-PPIN).

Several network topology features were selected from the CCS-PPIN: Average shortest path length, Betweenness centrality, Closeness centrality, Clustering coefficient, Degree, Eccentricity, Neighborhood connectivity, Number of directed edges, Radiality, Stress and Topological coefficient. The definition of these network features were shown in **Supplementary Table S3. Supplementary Table S4** offers the model topology features for the CCS-PPIN.

### Prediction model construction

Thirty-one diagnostic protein biomarkers collected from the CBD database were found on the CCS-PPIN. 31 non-DE proteins in the CCS-PPIN were randomly selected as the control group. We took 22 biomarkers and 22 non-DE proteins as the train set to establish an SVM model to predict biomarkers and another nine biomarkers and nine non-DE proteins as the test set to test the model performance. **Supplementary Material table S5** presents the dataset for Machine learning model construction.

A regression tree is an ML method that combines the advantage of a decision tree and regression. The aim of the regression tree is to find the best features and their cut off to classify the target. The regression tree implemented by R package “rpart” was utilized to choose the useful network features to distinguish the biomarkers from non-biomarkers.

SVM is a popular supervised machine learning method for classification issues. Using the selected features by regression tree, SVM was used to construct the topology model to predict new biomarkers in the CCS-PPIN, which was conducted by R package “kernlab”. We tried eight different kernels to train the SVM model to get the best prediction accuracy. Finally, Bessel kernel was chosen as the Kernel function in the SVM. A total of 2401 DE proteins in the CCS-PPIN was selected to predict new biomarkers. (**Supplementary Material table S6**)

### ROC test for the predicted biomarkers

The receiver operating characteristic (ROC) curve was used to identify the predicted biomarkers from the SVM model using the patients’ data provided by the GSE 44861 microarray data. The area under the Curve (AUC) of the ROC curve was recorded to compare the diagnostic accuracy of candidate biomarkers.

### PPI network and Biological function analysis

PPI network analysis, Gene Ontology (GO) annotation and KEGG pathway enrichment performed by String and Gluego on Cystoscope were conducted to analysis the candidate biomarkers calculated from our SVM model and confirmed biomarkers from the CBD database in biological interaction and function level.

### Multiple biomarkers identification

The predicted biomarkers were collected to combine as multiple biomarkers by logistic regression. ROC curve was drawn to test the diagnostic accuracy of multiple biomarkers.

## Results

**Figure 1** shows the analysis pipeline for this study.

**Figure 1.**
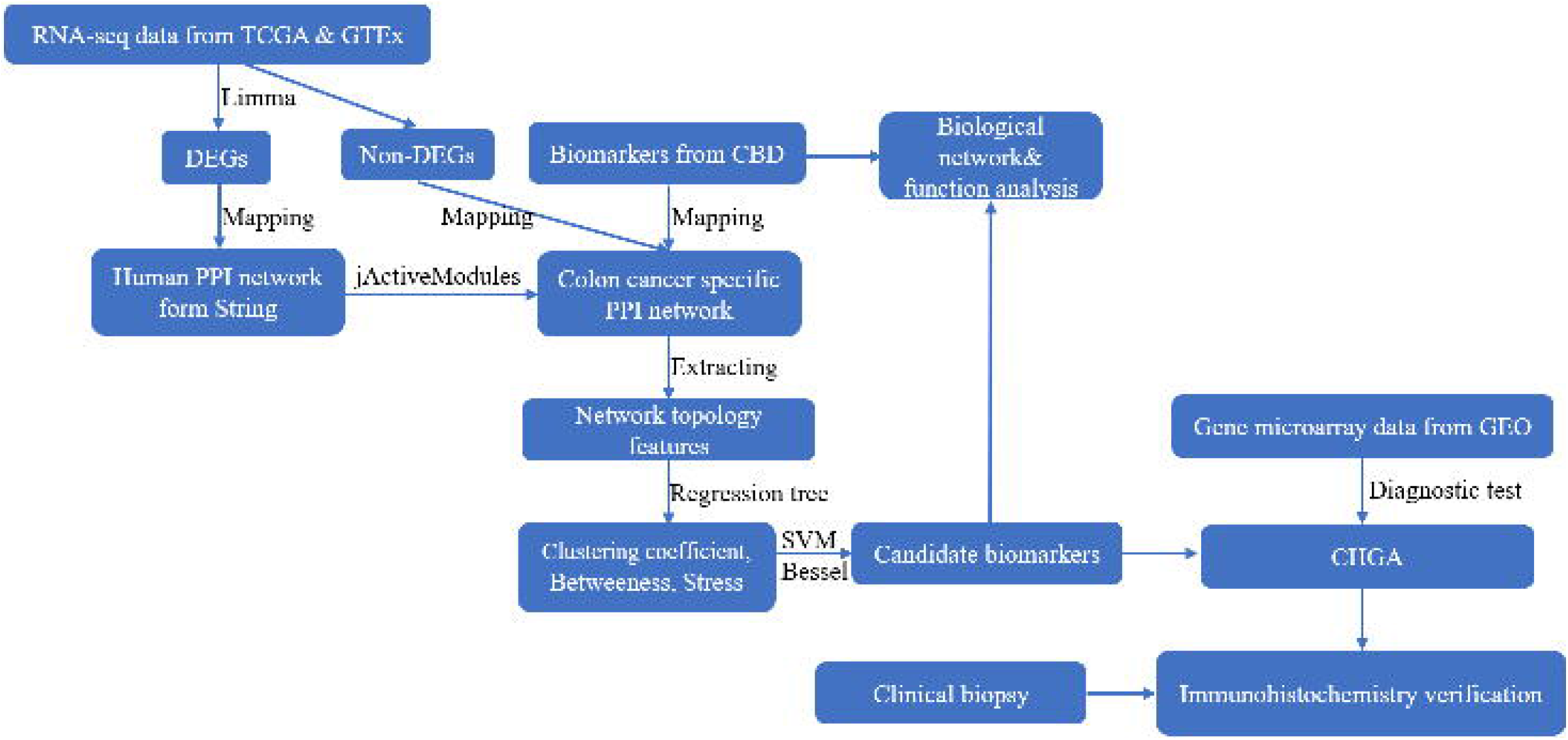
Schematic flow chart of the present study for early diagnosis of colon adenocarcinoma. The starting materials were derived from the RNA seq data in the TCGA and GTEx databases. Differential expressed analysis (DEA) between the colon cancer patients and normal controls was conducted. The differential expressed (DE) genes were then mapped to the Humman PPI network (from String) to construct colon cancer specific PPI network, and machine learning was used to predict new potential biomarkers based on the network features of the confirmed biomarkers from our CBD database. The diagnostic test (ROC test) of the predicted biomarkers were further verified in GEO microarray data. The candidate biomarker (CHGA) was finally confirmed by immunohistochemistry tissue microarrays.

### Colon cancer specific protein-protein interaction network (CCS-PPIN)

5562 colon cancer DEGs were identified based on the p-value and Log_2_FC. **Figure 2A** shows the DEGs position on Chromosomes. After mapping these DEGs using jActiveModules, we got the Colon cancer specific protein-protein interaction network (CCS-PPIN). CCS-PPIN contains 9624 nodes and 199553 edges. 11 original network topology features (Average shortest path length, Betweenness centrality, Closeness centrality, Clustering coefficient, Degree, Eccentricity, Neighborhood connectivity, Number of directed edges, Radiality, Stress, Topological coefficient) of each nodes were extracted from the CCS-PPIN.

**Figure 2.**
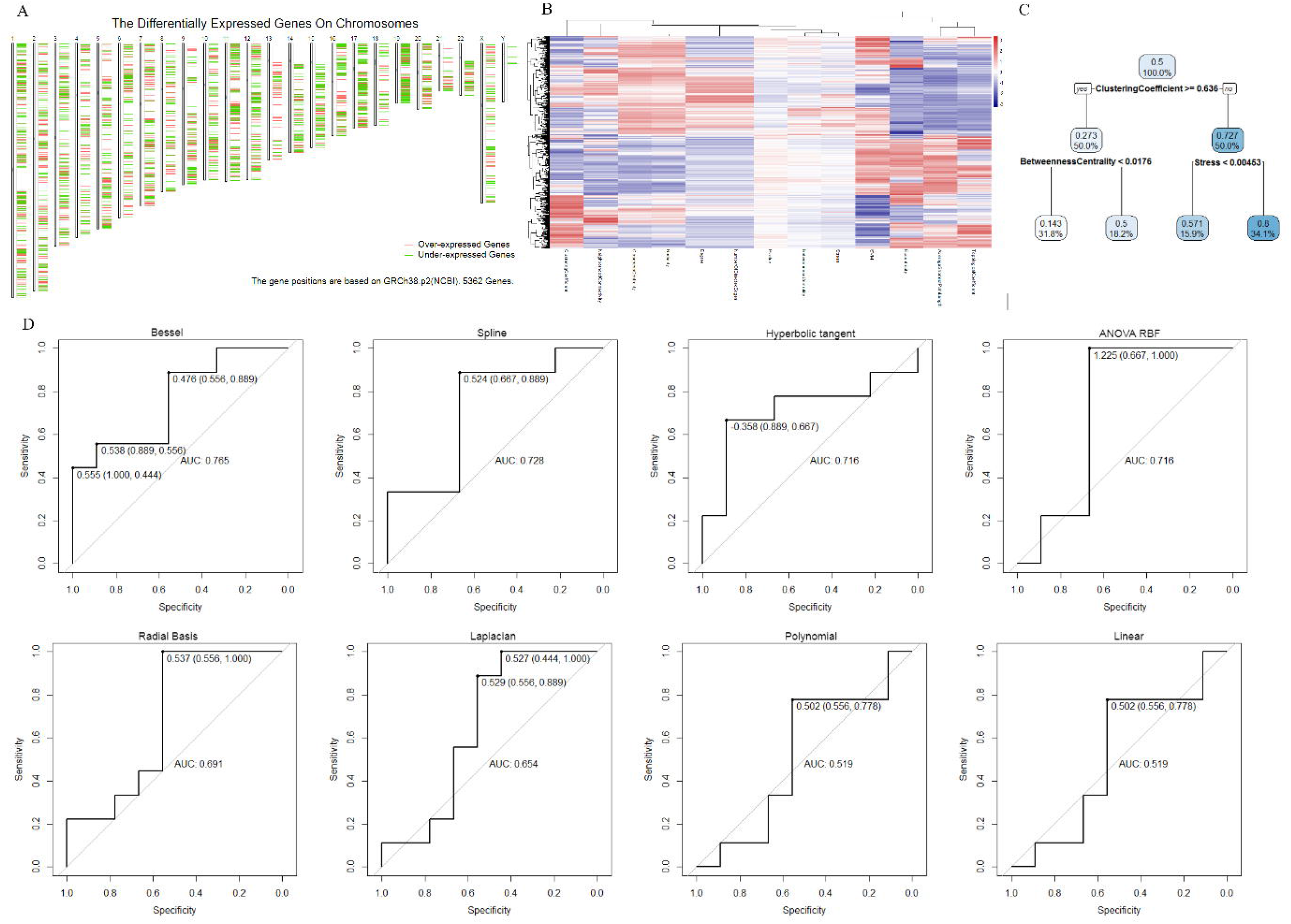
**A**. The differential expressed (DE) genes on various chromosomes. **B**. Regression tree in biomarker prediction model construction. Clustering coefficient, Betweenness centrality and Stress were selected as the features for the next SVM model. **C**. The results for ROC tests for different SVM models performed by different kennels. Bessel showed the best prediction accuracy. (AUC=0.765)

### Machine learning based biomarker prediction

A regression tree was conducted to select useful parameters for SVM model among the 11 original network features. Finally, Clustering coefficient, Betweenness centrality and Stress were selected. (**Figure 2B**)

We tried different kernels to train the SVM model using train data and predict the test data. ROC curve was selected to calculate the perdition accuracy. (**Figure 2C**) With its 0.765 prediction AUC, Bessel was selected as the kernel for the final biomarker prediction SVM model. Figure 2D shows the There were 2401 DE proteins on the CCS-PPI, which was selected to predict new colon cancer biomarkers using the SVM model. Through the model, each protein will be given a point, which is the possibility to be a biomarker. We set 0.99 as a cut off for the SVM point, and **Figure 3A** presents the 12 predicted biomarkers.

**Figure 3.**
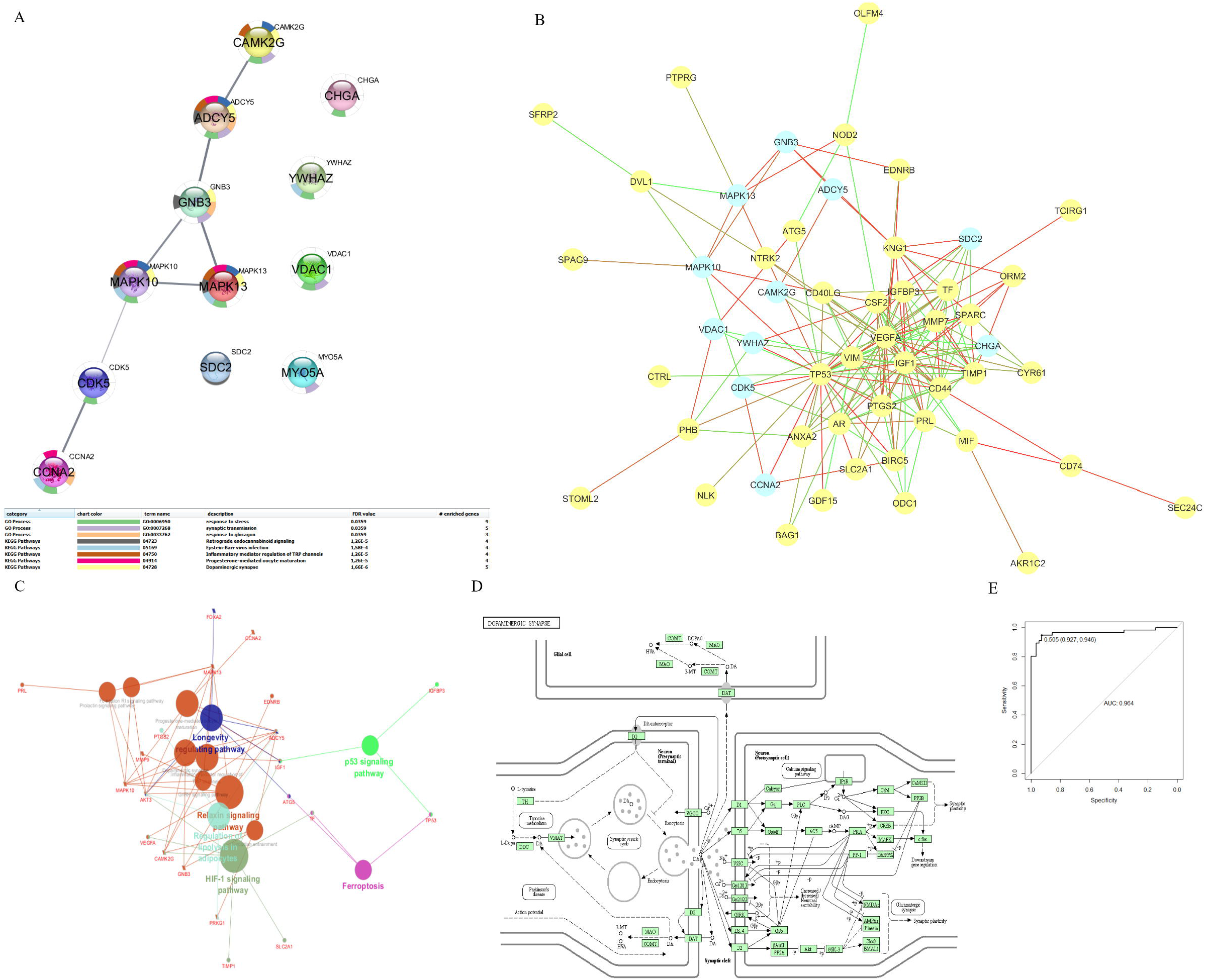
PPI network and biological function analysis of the predicted biomarkers. A. The candidate biomarkers that had a strong relationship were mapped in the same pathway. **B**. PPI network for predicted and confirmed the biomarkers. Generally, the predicted and confirmed biomarkers have strong relationships with each other; specifically, not as the other ten predicted, which are connected closely, CHGA and SDC2 are separated from them and are hubs in their belonged small networks. **C**. KEGG pathway enrichment analysis to predict and confirm biomarkers. There are some overlapping pathways among confirmed and predicted biomarkers, and 5 of them were mapped on the Dopaminergic synapse pathway. **D**. Dopaminergic synapse pathway. E. ROC curve for multiple biomarkers.

### Verification of predicted biomarkers

ROC analysis performed by gene expression data were used to test the diagnostic value of candidate biomarkers predicted by the SVM model. 11 predicted biomarkers were found on the GPL, and they all showed high AUC (bigger than 0.5). Among them, CHGA has the best AUC (0.9). **Supplementary Table S6** listed all the tested proteins (DEGs) along with their SVM point and diagnostic AUC.

Scatter plots and box plots of network features on predict model for predicted biomarkers, and other genes are shown in **Figure 4**. Significant differences were identified among predicted/identified biomarkers and other genes.

**Figure 4.**
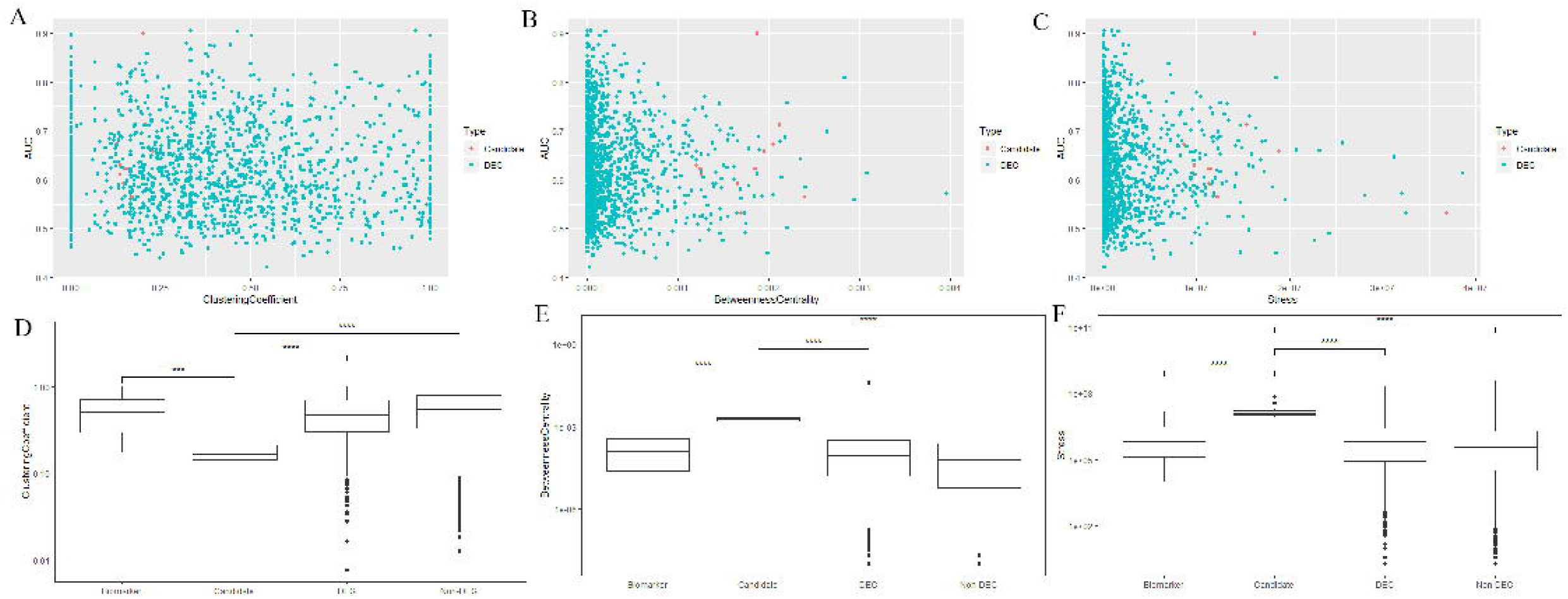
Comparison of network features of predicted biomarker candidates with other genes. A-C: Cluster coefficient (A), Betweenness centrality (B), and Stress (C) of candidate biomarkers (red points) and DEGs (green points) on CCS-PPIN. Biomarker candidates showed specific features in Cluster coefficient (0.125-0.25) and Betweenness centrality (0.001-0.0025). D-F: Box plot of Cluster coefficient (D), Betweenness centrality (E), and Stress (F) of identified biomarkers, biomarker candidates, DEGs and non-DEGs. Biomarker candidates showed significant differences with other genes.

### PPI network and biological function analysis for predicted biomarkers

We used the String database to explore the relationship between these predicted biomarkers in PPI network and biological pathways. **Figure 3A** showed the PPI network and biological function analysis result of 12 predicted biomarkers. CCNA2, CDK5, MAPK10, MAPK13, GNB3, ADCY5 and CAMK2G showed a strong relationship in the PPI network. KEGG pathway enrichment analysis showed that 5 of them were mapped on the Dopaminergic synapse pathway. (**Figure 3D**) According to the GO annotation, 9 of these predicted biomarkers were related to the Response to stress.

### Relationship for reported and predicted biomarkers on PPI network and biological function

In order to investigate the relationship between already reported biomarkers from the CBD database and newly predicated biomarkers, we mapped them together in the human PPI network. (**Figure 3B**) And we found that most of predicted biomarkers were the close neighbourhood of confirmed ones, and some famous biomarkers such as TP53, VEGFA, and IGF1 were still hubs for this PPI. 10 predicted biomarkers had direct relationships with each other but not SDC2 or CHGA, which occupied two separate positions beside others. What’s more, from Table 1, we found that SC2 and CHGA had the highest AUC (0.71 and 0.90) on the ROC curve of the diagnostic test.

We performed the KEGG pathway enrichment analysis for the confirmed and predicted biomarkers and mapped together with the results in **Figure 3C**. There were two overlapping for the two group biomarkers: Inflammatory mediator regulation of TRP channels, Progesterone-mediated oocyte maturation. p53 signalling pathway and Ferroptosis were the two most confirmed biomarkers mapped pathways, and GnRH signalling pathway was the most mapped pathway for only predicted biomarkers.

GO annotation in Biological process, Cellular component, Immune system process and Molecular function level were conducted **(SM Table S7)**, and we found three overlapping pathways for the confirmed and predicted biomarkers: Positive regulation of osteoblast differentiation, Morphogenesis of an epithelial sheet, Positive regulation of fibroblast proliferation and Regulation of fibroblast proliferation. CCNA2, as a predicted biomarker, was mapped on all of these 4 pathways.

### Identification of Multiple biomarker

We combined the predicted biomarkers as multiple biomarkers via logistic regression and using AUC analysis to test its diagnostic value. The ROC on the AUC curve of multiple biomarkers is 0.964. **(Figure 3E)**

### Verification for CHGA

With its best performance in a diagnostic test, CHGA was selected to make further verification. **Figure 5** presented the IHC result for CHGA in CRC. CHGA protein was positively expressed in the normal colon and adjacent colon mucosa (the brown colour) and lost the CHGA expression in the colon cancers regardless of well, moderate or poor differentiation of the cancers.

**Figure 5.**
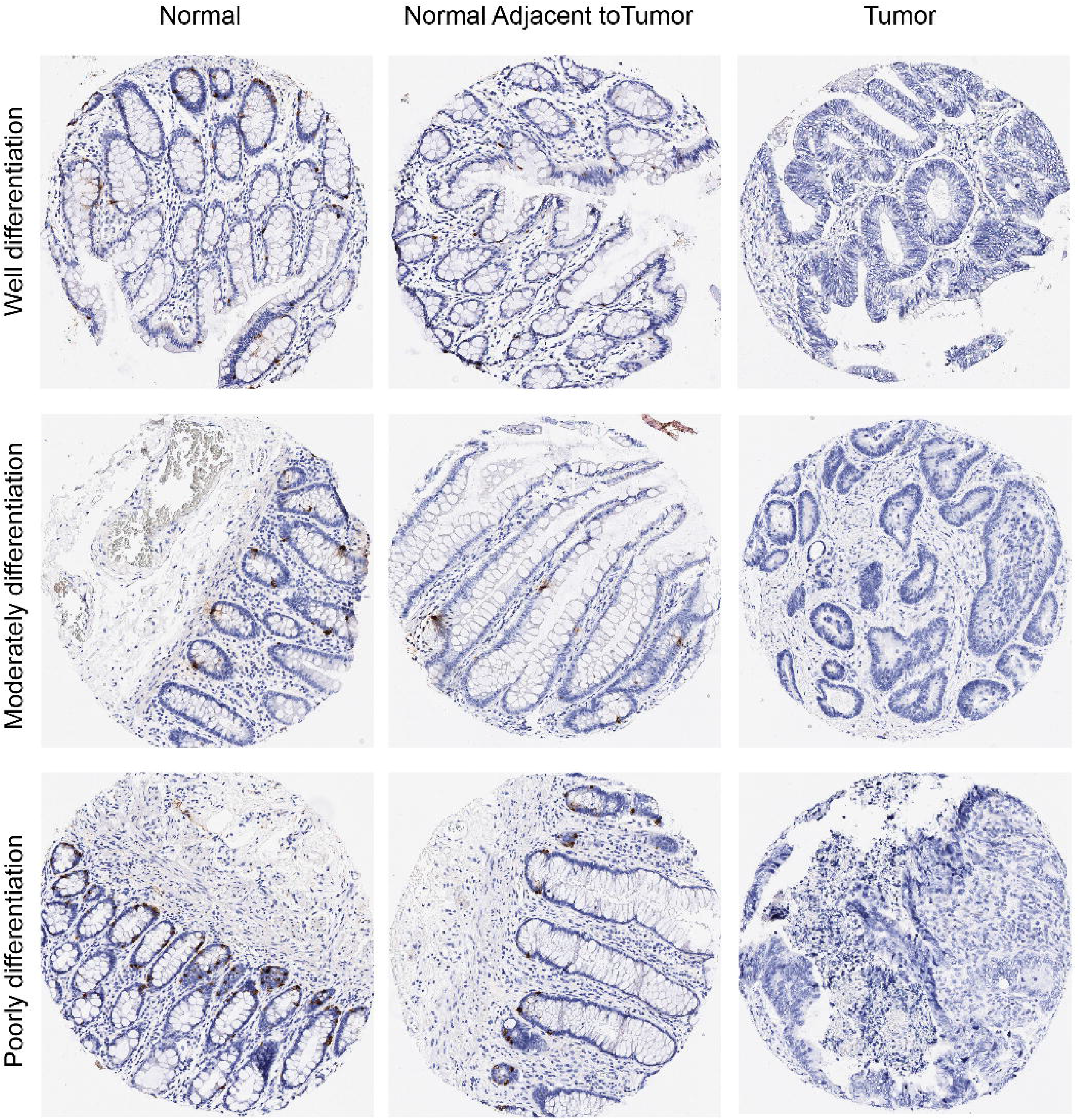
Protein expressions of CHGA in normal colon mucosa, adjacent mucosa and colon cancers from the same patients. CHGA protein was positively expressed in the normal colon, and adjacent colon mucosa (the brown color) and absolutely lost the CHGA expression in the colon cancers regardless of well, moderate or poor differentiation of the cancers. Magnifications 10x and 40x.

Figure 6 showed the expression of CHGA in normal and cancer patients (Figure 6A: colon cancer patients, 6D: rectal cancer patients, 6G: CRC patients), diagnostic ROC tests (Figure 6B: colon cancer patients, 6E: rectal cancer patients, 6H: CRC patients), and survival tests (Figure 6C: colon cancer patients, 6F: rectal cancer patients, 6I: CRC patients). CHGA showed significantly lower expression in CRC patients than normal controls and behaved well in the diagnostic test. (AUC: 0.995) However, CHGA may not be served as a prognostic biomarker for CRC. (p-value on survival test: 0.24, 0.38, and 0.13)

**Figure 6.**
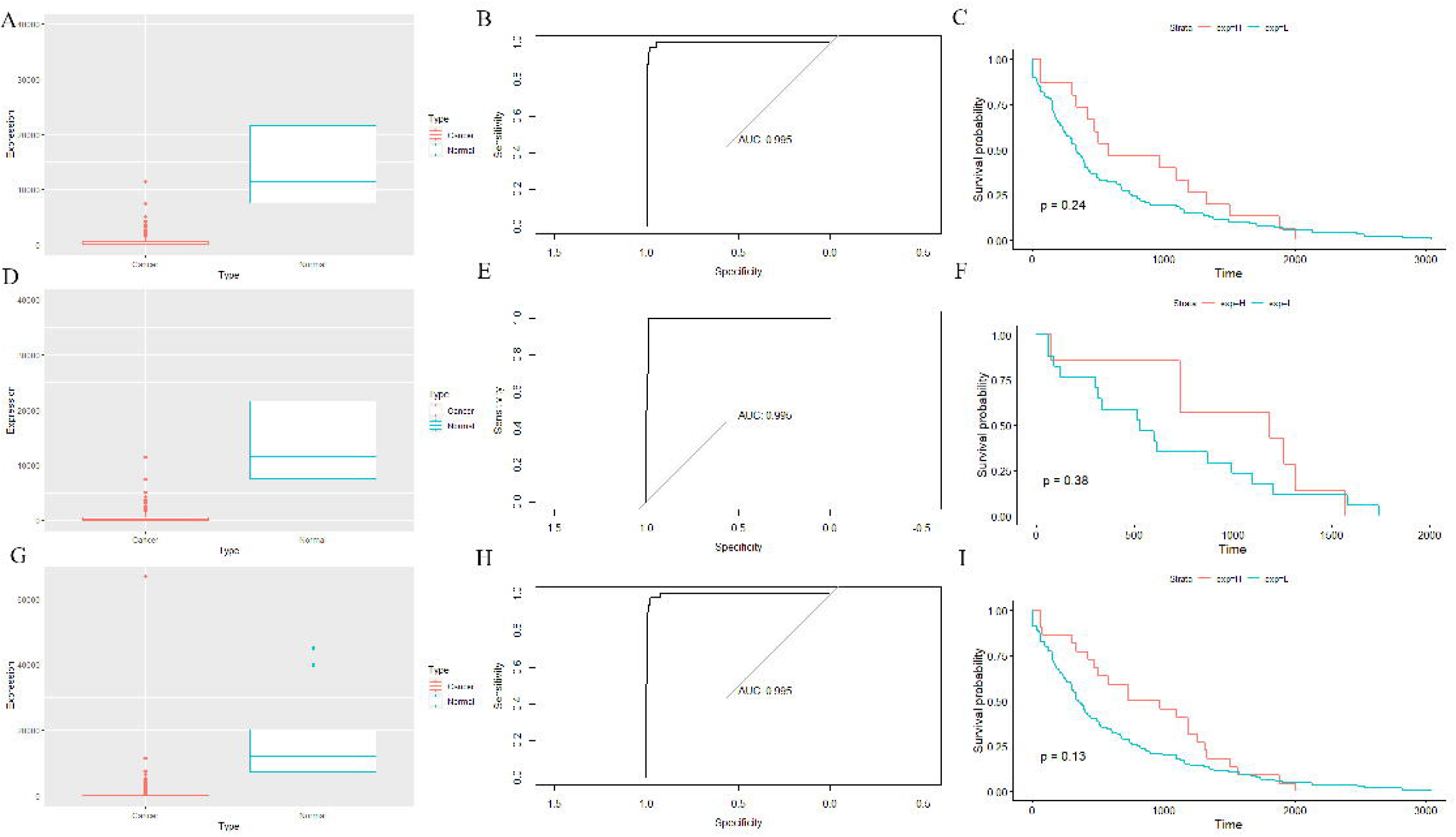
CHGA expression distribution of colon cancer tissues (A), rectal cancer tissues (D) and CRC (G) compared with normal controls. CHGA showed a significant difference in cancer tissues with normal controls. Diagnostic ROC test for CHGA in colon cancer (B), rectal cancer (E) and CRC (H). With AUCs of 0.995, GHGA showed a high potential of being a good diagnostic biomarker in CRC. Survival curves of CHGA in colon cancer patients (C), rectal cancer patients (F), and CRC patients (I). CHGA performed poor in the prognosis of CRC. (p value=0.24, 0.38, and 0.13)

## Discussion

Colon cancer is one of the most common types of cancer [31]. The detection of new biomarkers is extremely important in the diagnosis of colon cancer [5]. Colonoscopy has been considered a golden test for colon cancer diagnosis [3]. However, it is invasive and expensive. As such, the development of other diagnosis methods is still needed. Biomarkers have been proven with the ability to provide useful additional evidence in colon cancer diagnosis [32]. However, only a few biomarkers have been used in clinical. Hence, the detection of new biomarkers is extremely needed.

As mentioned in the introduction, both dry and wet experiments have their significant advantage. However, dry experiments are always doubted with their false positives, and wet experiments are limited by their lab environments. Hence, more and more scientists suggest combining the wet and dry experiments together, by which to make the results of studies more comprehensive and credible. In this study, we used two ML methods (Regression tree and SVM) to construct the biomarker prediction model based on the PPI network topology features and predicted CHGA as a novel diagnostic biomarker, which was further verified by IHC. A regression tree was used to find the best features on the PPI network, which were selected as the final features for the SVM prediction model. The kernel is an essential part of SVM. We tried eight different kernels in the SVM prediction model and tested their prediction accuracy using the ROC test. Finally, the “Bessel” kernel was selected with its 0.765 AUC.

Recently, many bioinformatics studies used ML algorithms to predict new biomarkers [22, 23]. Compared with previous studies, our present study used all the reported colon cancer biomarkers collected from our CBD database as train data to predict new biomarkers, which increased the credibility. Furthermore, prediction features were selected from a human PPI network optimized by jActiveModules, which increased the robustness. We used network topology features on the PPI network as prediction features. Compared with biological features, topology features can decrease the negative influences caused by sample heterogeneity and size for the predicted model [33]. There are two predicted biomarkers (CHGA and SCD2) that performed best in the diagnosis test. Interestingly, unlike the other ten predicted biomarkers connecting with each other, CHGA and SCD2 occupy independent positions on the PPI network. Furthermore, CHGA and SCD2 are both hubs on this PPI network, and they are close to the core networks of identified biomarkers. Many studies convince that biological networks share similar features with the human social network. CHGA and SCD2 are just like heroes in the social network: they are alone but inflect many other points in their small networks. So we predict that some palmary biomarkers share a similar network position in biological networks as heroes in social networks, and we call them “hero biomarkers”.

We have identified the diagnosis value of CHGA as a biomarker in colon cancer using meta-analysis [17]. In this study, IHC was further used to verify the diagnosis value for CHGA in colon cancer, which proved that CHGA could be a promising biomarker in biopsy (Figure 4).

Biological functional analysis has been conducted to verify the prediction results. We found some overlapping enriched pathways for the predicted and reported biomarkers, which convinced our results. Meanwhile, the results of biological function analysis inspire researchers to detect new biomarkers in these pathways.

Multiple biomarkers combined by several single biomarkers have been convinced to improve the diagnosis effect in many previous studies [7, 18]. In the present study, we combined the 12 predicted biomarkers as multiple biomarkers and found that they showed significantly high diagnosis accuracy. Hence, we recommend that multiple biomarkers could be used further in the clinical train.

## Conclusion

We used ML to predict new biomarkers for colon cancer diagnosis based on the PPI network and found 12 candidate biomarkers, of which CHGA showed good diagnostic performance in gene expression data and IHC. We combined these predicted biomarkers as multiple biomarkers and showed better performance than solely. Further, these predicted biomarkers share some pathways with reported biomarkers, and these pathways may be pivotal pathways for further biomarker discovery for colon cancer. We also tested the diagnosis value of CHGA in rectal cancer and showed promising results. In conclusion, CHGA may be a future diagnosis biomarker for CRC patients.

## Data Availability

All data produced in the present study are available upon reasonable request to the authors

## Acknowledgments

The authors are grateful to the staff in our research groups who were involved in the study for their valuable contributions and discussions. The authors would like to express their gratitude to Siyu Qiao and Yu Shao for their help in network topology knowledge and Guang Hu, Dirk Repsilber, Xuye Yuan and Shunming Liu in Machine learning.

